# Depression following traumatic brain injury is characterised by dominant and recurring brain loops in self-referential areas

**DOI:** 10.1101/2020.08.14.20175000

**Authors:** Ioannis Pappas, Laura Moreno-López, Ellen L. Carroll, Anne Manktelow, Joanne G. Outtrim, Jonathan P. Coles, Virginia F. Newcombe, Barbara J. Sahakian, David K. Menon, Emmanuel A. Stamatakis

## Abstract

Depression is a major albeit neglected complication in patients with traumatic brain injury (TBI). Elucidating its neural correlates remains an important milestone with respect to understanding the disorder and helping with the rehabilitation process. Towards this direction, neuropsychological theories have proposed abnormal brain dynamics as the neural basis of depressive symptomatology. This observational study addressed the question of whether depression in TBI patients is related to abnormal brain dynamics using a sample of 81 TBI patients with depressive symptomatology. To explore brain dynamics we employed the Hidden Markov model that utilises resting-state fMRI data to identify the states that the brain visits sequentially during scanning. Spatial (highest activated regions) and temporal (occupancy, switching rate) characteristics of these states were used to analyse the networks involved and probe differences between depressed and non-depressed TBI patients. We found a significant positive association between depression score and the fractional occupancy and switching rate of two specific states that distinguished between depressed and non-depressed TBI patients. These states spanned default mode, subcortical and cerebellar regions while also forming a temporally coherent “metastate” that the depressed brain would recurrently visit. Depression in TBI patients is characterised by abnormal recruitment and repetitive sequencing between certain neural networks. These results point to the existence of a reinforced, self-referential circuitry that could provide the basis for targeted therapies during the recovery process.

## Introduction

Traumatic brain injury (TBI) is a major public health issue with approximately 50-60 million of new cases every year (1). One of the most important, albeit largely neglected, complications. following TBI is the development of major depressive disorder (2). Depression following TBI has devastating consequences as it increases the risk of suicide, impairs overall quality of life, and affects interpersonal, occupational, and social functioning (3). From a neuropsychological perspective, depression has been associated with abnormal emotional regulation and information processing suggesting that this disorder might be related to the dysfunction of an extensive network of regions (4,5). From a neuroanatomical perspective, TBI causes diffuse or focal patterns of brain damage that may contribute to the depressive symptomatology (6)

Neuroimaging techniques such as resting-state functional MRI can now reliably map the functional architecture of the brain (i.e. functional connectivity), by measuring the correlations between blood oxygen level dependent signals from different brain regions usually over the whole duration of the resting-state scan (static functional connectivity)(7). Studies using static, resting-state functional connectivity have shown that depression might be associated with the aberrant functional connectivity of brain systems or networks, mainly the default mode network (DMN)(8,9), the frontoparietal network including the dorsolateral prefrontal cortex/DLPFC) and the salience network (including the insula and anterior cingulate cortex/ACC) associated with the regulation of emotional processing (10).

Posited theories regarding brain dynamics during depression go above and beyond static functional connectivity and emphasise the dynamic recruitment and switching between different networks (11). This scheme is self-reinforcing, meaning that there is ongoing switching between different networks that maintains the depressive state potentially via deregulated bottom-up and top-down control from and to the DLPFC (12,13). Thus it is possible that depressive symptoms post TBI relate to abnormal temporal processing and that this prevents these patients from planning and conducting daily actions.

Towards this direction a growing body of literature has used tools that quantify time-varying functional connectivity (TVFC) (14) in order to characterise the fluctuating recruitment of these networks in patients with depression (15,16). Although these studies provide valuable pieces of information about brain dynamics during depression, they have several limitations. The most important limitation is that standard TVFC methods quantify the temporal variability of functional connectivity over windows of time thus being insensitive to brain transitions happening at a faster scale (17,18).

This calls for increased temporal and spatial specificity in the exploration of brain dynamics in TBI depression. In this study, we used a completely data driven method for identifying dynamics in the depressed brain post TBI. We used the Hidden Markov Model HMM (18,19) in a large cohort of 81 patients post TBI with and without depressive symptomatology. Briefly, this method characterises, at a temporal resolution as high as the modality allows, brain dynamics by looking at the transitions between spatially distinct networks or states that the brain makes sequentially during scanning. to test two hypotheses. Using this method we investigated two hypotheses: First, we assumed that TBI patients with depressive symptoms would be characterised by an abnormal recruitment and switching rate of specific states compared to nondepressed patients. In addition, we assumed that these would form a metastate in the sense that the brain would display a preference for cycling between these states in line with previous theories discussing circuitry implicated in the self-reinforcing of the depressive symptoms. Second, we hypothesised that these brain states would represent networks responsible for abnormal regulation of information as it has been previously proposed by neuropsychological models of depression.

## Results

### 2.1 Hidden Markov model in the patient cohort

Our first question was “Are brain dynamics of TBI patients associated with recurrence of specific states?” To answer this question, we calculated state time courses by applying the HMM to concatenated data from 81 individuals with TBI and showed varying depression scores (see Methods). An advantage for using one model for both TBI depressed and non-depressed individuals is that it allows us to derive states common to both groups thus making the exploration of activity spatio-temporal characteristics meaningful between the two groups. We then calculated the average time spent in each state-namely their fractional occupancy/FO. Several states showed high fractional occupancy while other states were rarely occupied (**Fig. 2a**).

**Figure 1.**
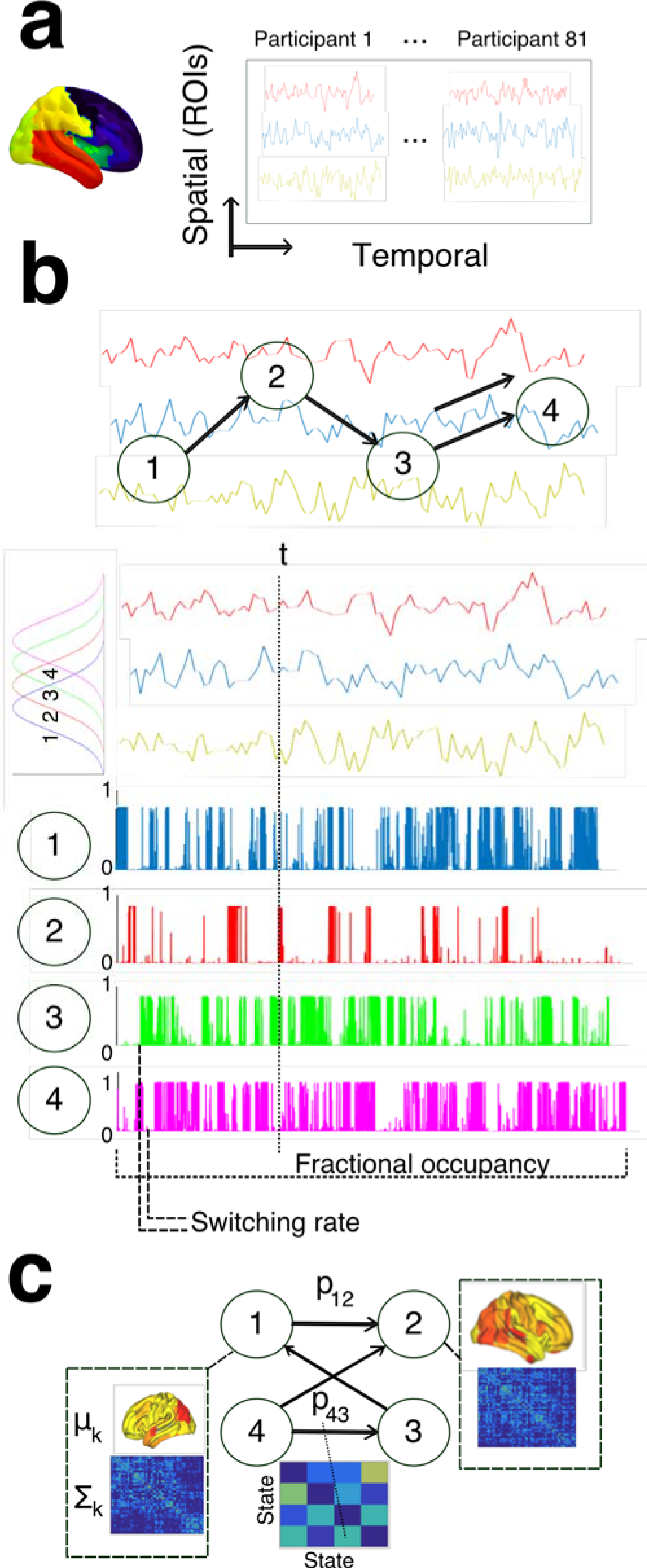
Deriving brain dynamics from the patients’ resting-state data using the Hidden Markov model (HMM) **a)** Time courses for different brains regions of interest (ROIs) were obtained for each participant using a specific parcellation. Time courses were then concatenated across participants in the temporal dimension. **b)** HMM assumes that brain activity is organized into discrete states that the brain occupies and switches between at each time point of the scanning. Each time point of brain activity is characterised by a mixture of multivariate Gaussian distribution describing the probability of the brain being active in each state. **c)** The HMM was applied to the concatenated ROI data and resulted in a number of states each one characterised by a mean (activity) and covariance ((connectivity)). Fractional occupancy/FO of each state was defined as the mean probability of that state being active over the duration of the scanning. Switching rate was defined as the difference in their activation probabilities between consecutive time points.

**Figure 2.**
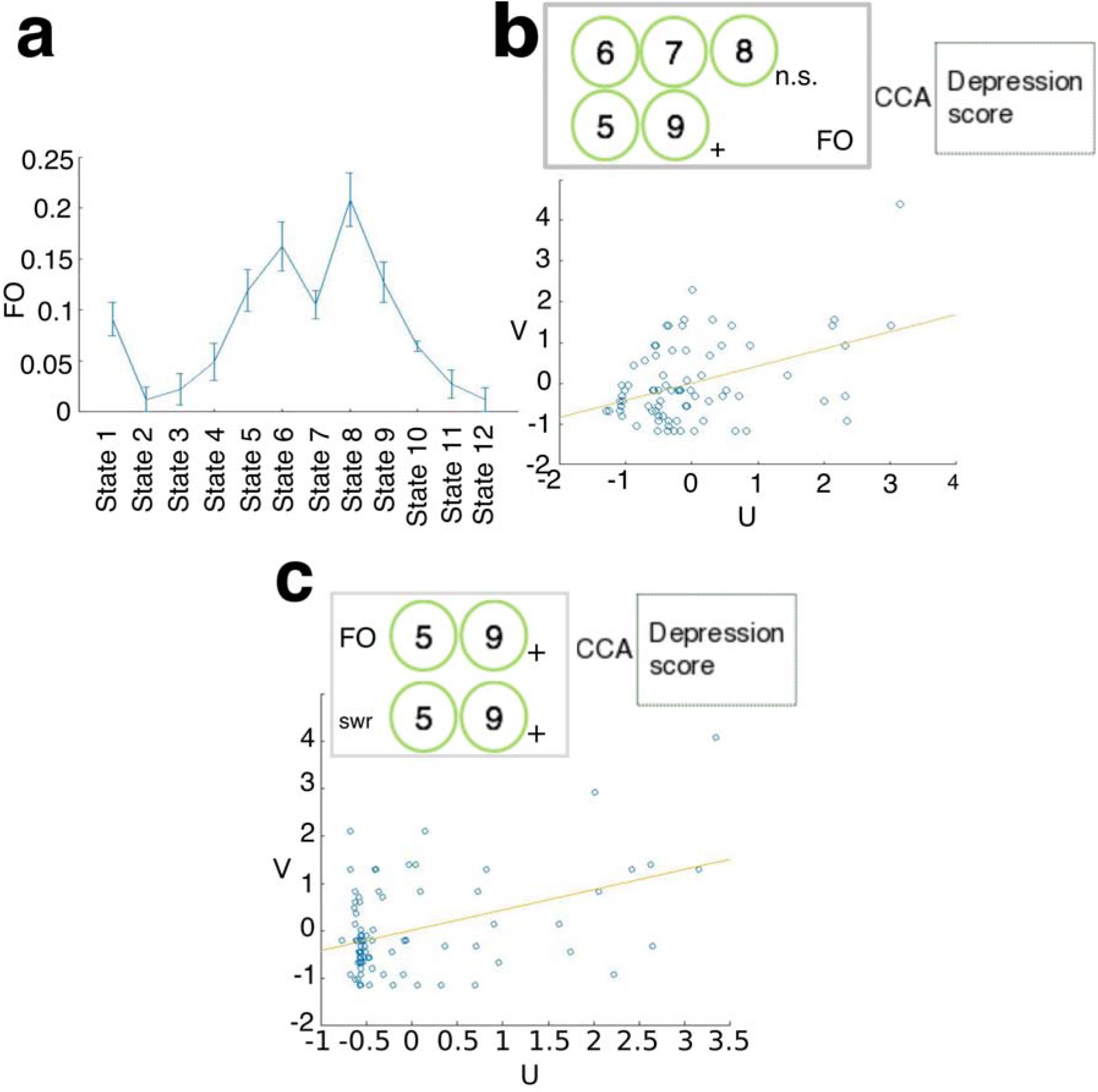
Correlations between depression and the fractional occupancy and switching rate of brain states. **a)** We ran HMM on the patient cohort and it resulted in five states with highest FO (average probability of them being active during the scanning). **b)** Using CCA we identified a significant mode of co-variation (CCA mode as characterised by the variables U and V each one representing linear combinations of predictor and predicted variables respectively) between their FOs and the depression score. States 5 and 9 were the only ones that positively correlated *(post hoc)* with this CCA component and we used their FOs as predictors in a second CCA to predict the participants’ depression scores. **c)** The FO of states 5 and 9 and their switching rate (swr) was included in a secondary analysis to produce a CCA component that was also positively correlated with the depression scores. Both FOs and the switching rate were positively correlated (post hoc) with this CCA mode. Vertical lines in the FO plot show standard error of the mean. (+) symbols indicate the positive *post hoc* correlation of variables with the CCA mode. n.s. indicates non-significant *post hoc* correlation of variables with the CCA mode.

We chose the FO of the top five occupied states as predictor variables in a CCA in order to see if there was a significant mode of co-variation (or CCA component) between their FOs and depression scores. The reason for choosing these states were a) due to their high increased occupancy > 0.1 across patients (permutation testing p-value uncertainty for hypothesis FO<=0.1 [0.09 0.13] [0.9 0.93] [0.72 0.77] [0.92 0.95] [0.08 0.12] respectively for each state) b) to avoid excessive multiple comparisons issues and c) to enhance interpretation. We found a significant CCA mode between the FO of these states and the participants’ depression scores meaning high recurrence in these 5 states were associated with high depression symptoms (r=0.4278, p=0.0083, corrected for multiple comparisons across all modes estimated, **Fig. 2b**). Calculating this CCA mode over a subset/training set of the data and testing on the remaining/test set of the data (mean correlation on test set over 1,000 random 80%-20% train-test splits r=0.3412) showed that the relationship between FO and depression was not a statistical property of a subset of the data but it could be generalised to the entire TBI cohort.

Correlating *post hoc* each state’s FO with the CCA mode gives a weight of the contribution of each variable to the mode of covariance. To that end, we found that out of the top five high occupancy states only states 5 and 9 were positively correlated to the CCA mode (r=0.76, p<0.001, r=0.91,p<0.001, Bonferroni corrected, states 6,7,8 r=-0.0599,r=0.1093,r=-0.052).

This result indicates that these two states were contributing more to the variance in the participants’ depression scores than the other high occupancy states. These results (a significant CCA mode with two prominent states showing a relationship with the depression score) were verified using different parameters of the model (**Supplementary Figure 2**) as well as a different parcellation (**Supplementary Figure 3**).

Next, we asked whether the switching rate of states 5 and 9 would also relate to depression on the basis that the high temporal variability in the recruitment of each state also contributes to the depressive symptomatology. We used the FOs of states 5 and 9 alongside their switching rate to predict depression scores in a separate CCA. We found a significant CCA mode with (r=0.4328, p=0.0030, corrected for multiple comparisons across all modes estimated, **Fig. 2c**) with both FOs of states 5 and 9 (r=0.5656 p<0.001, r=0.8404 p<0.001, Bonferroni corrected), and switching rate (r=0.6911,p<0.001, Bonferroni corrected) strongly correlated *post hoc* with the CCA mode. Additional train/test evaluations showed high correlation between FO of states 5 and 9, their switching rate and depression score (mean correlation over 1,000 random 80%-20% train-test splits r=0.2812).

### 2.2 Spatial and temporal characteristics of states 5 and 9

Given their prominence in predicting depression scores in the TBI cohort, we sought to spatially characterise these states (see Methods). We found that state 5 included mostly brainstem, cerebellar regions, most of the thalamus and cingulate cortex while state 9 included mostly the precuneus and angular gyri, regions of the brain that are known to be part of the DMN (**Fig. 3**). Thus high recurrence and switching of states 5 and 9 would include recurrent activation of subcortical and default mode regions that have been associated with mood regulation and internal cognitive function (35,36).

**Figure 3.**
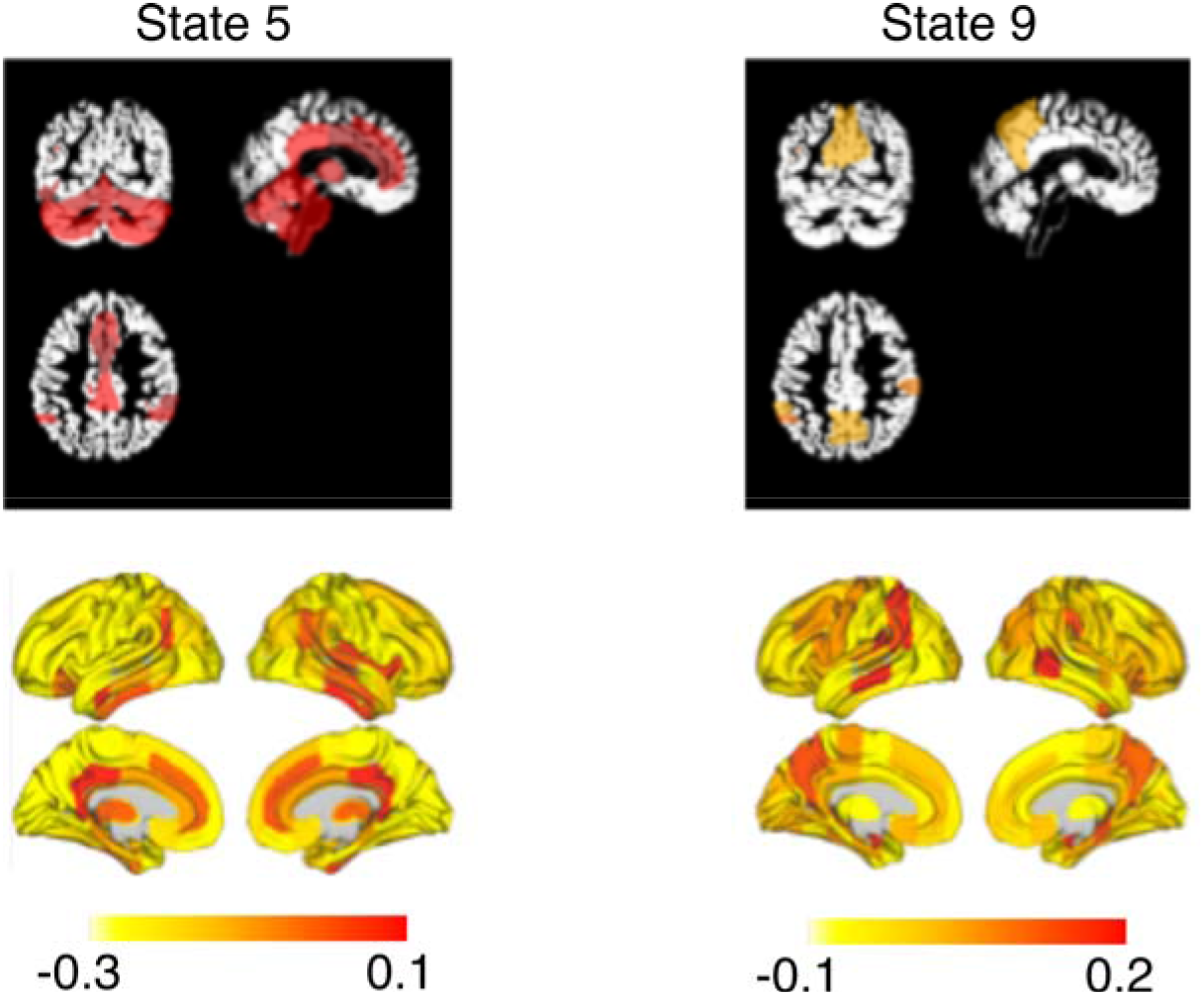
Spatial characteristics of states 5 and 9. Using the PCA loadings we projected the states’ 5 and 9 activity back into standard space. Each pair of panels shows axial, sagittal and coronal views of the regions with the highest activity (defined as the regions with the top 40% of activity), as well as full heat maps with the activity of all the ROIs. Numbers in the colorbars reflect the level of activation for each region in its respective state.

### 2.3 Difference in brain dynamics between depressed and non-depressed TBI patients

We next asked whether the previously found brain states were evident of the patients’ level of depression and differed between patients with high and low depression scores. Thus we investigated differences in the temporal characteristics of states 5 and 9 (FO and switching rate) between the TBI depression and TBI non-depression cohorts. We found a significantly higher FO (z=3.1681, p=0.0015, z=2.92, p=0.0035) and switching rate between states 5 and 9 (z=3.2094, p=0.0013) in the TBI depression group compared to the TBI no depression group (**Fig. 4a, 4b, 4c**). Collectively patients that had high depression score showed higher recruitment and switching in states 5 and 9. In conjunction with our previous observation regarding the spatial features of these states, this result suggests that depressive symptomology is characterised by high recruitment and recurrence of self-referential-related circuitry (5). It is worth noting that the mean grey matter intensity was not different between groups (two-tailed two-sample t-test, p=0.6847) thus the observed difference in their dynamics could not have been due to structural abnormalities that previous depression studies have suggested as an important marker for depression (37).

**Figure 4.**
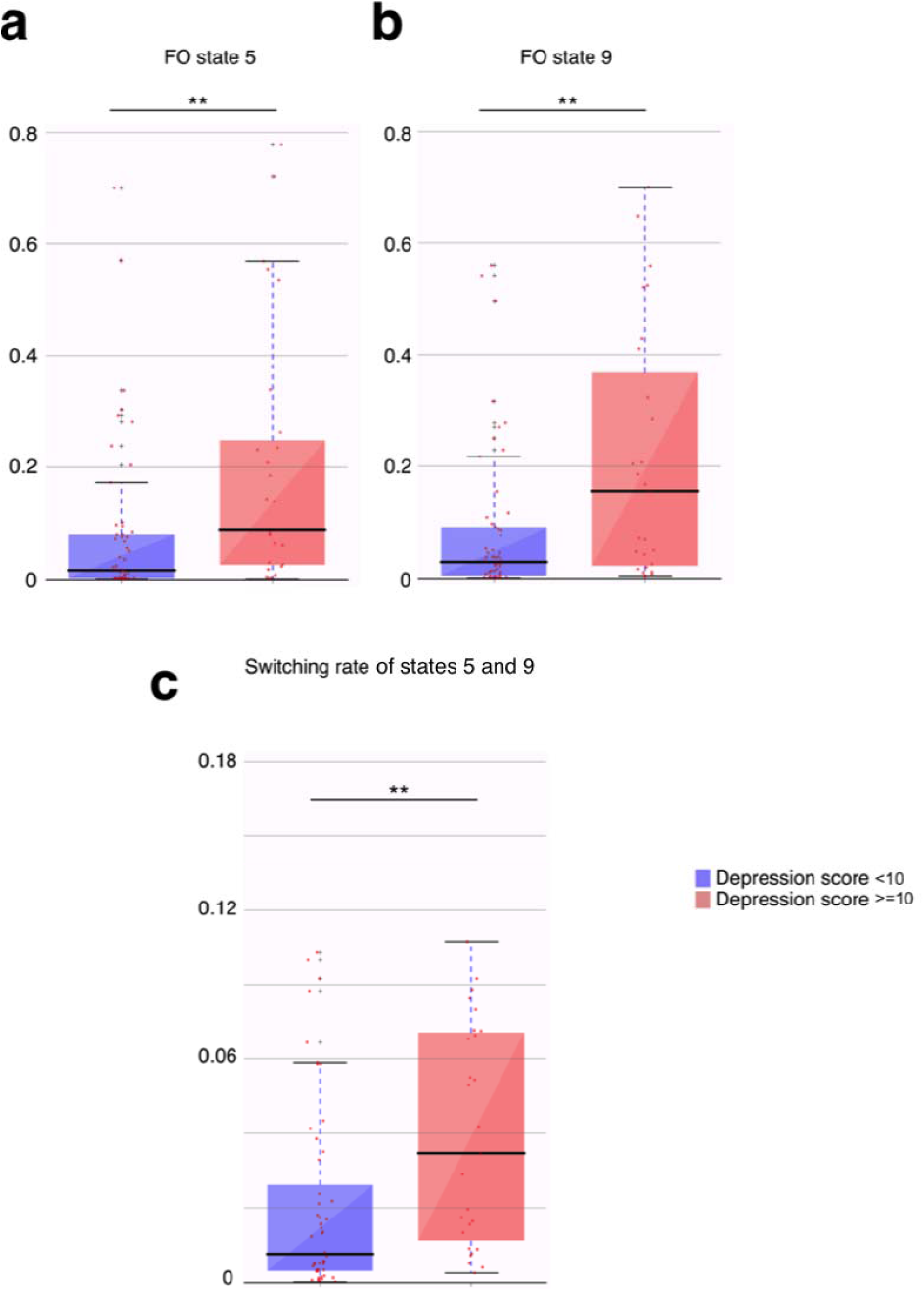
Fractional occupancy and switching rate between states 5 and 9 were higher in the TBI patients with high depression scores compared to those with low depression scores. The patient cohort was divided based on whether their BDI depression score was less than 10 (blue) or greater and equal than 10 (red). Panel **a** shows the FO differences between these two groups for state 5. Panel **b** shows similar results for state 9. Panel **c** shows the differences between these two groups in terms of the switching rate of states 5 and 9. Boxplots’ thick lines show median values and whiskers represent 1.5 times the inter-fourth range. ** stands for p < 0.01 significance.

Previous neuropsychological models have emphasised the role of self-reinforcing negative schemata, potentially produced by ongoing temporal deregulation between limbic and executive control regions (11). In light of this we wanted to see if the two states that were most prominent for depression were temporally related with the assumption being that the brain would preferentially cycle between these sates and not in other states. Results on the patient cohort revealed that the FOs of states 5 and 9 clustered together suggesting their high similarity (**Fig. 5a**). With respect to the switching rate, we found that HMM would switch significantly between states 5 and 9 indicating that when brain dynamics occupied state 5 or 9 there was a high possibility of cycling between these states rather than switching to another state (**Fig. 5b**). These two different methodologies converged to the conclusion that the temporal characteristics of states 5 and 9 were highly interrelated in the patient cohort suggesting that their temporal properties form a complementary mechanism for the depressive symptoms.

**Figure 5.**
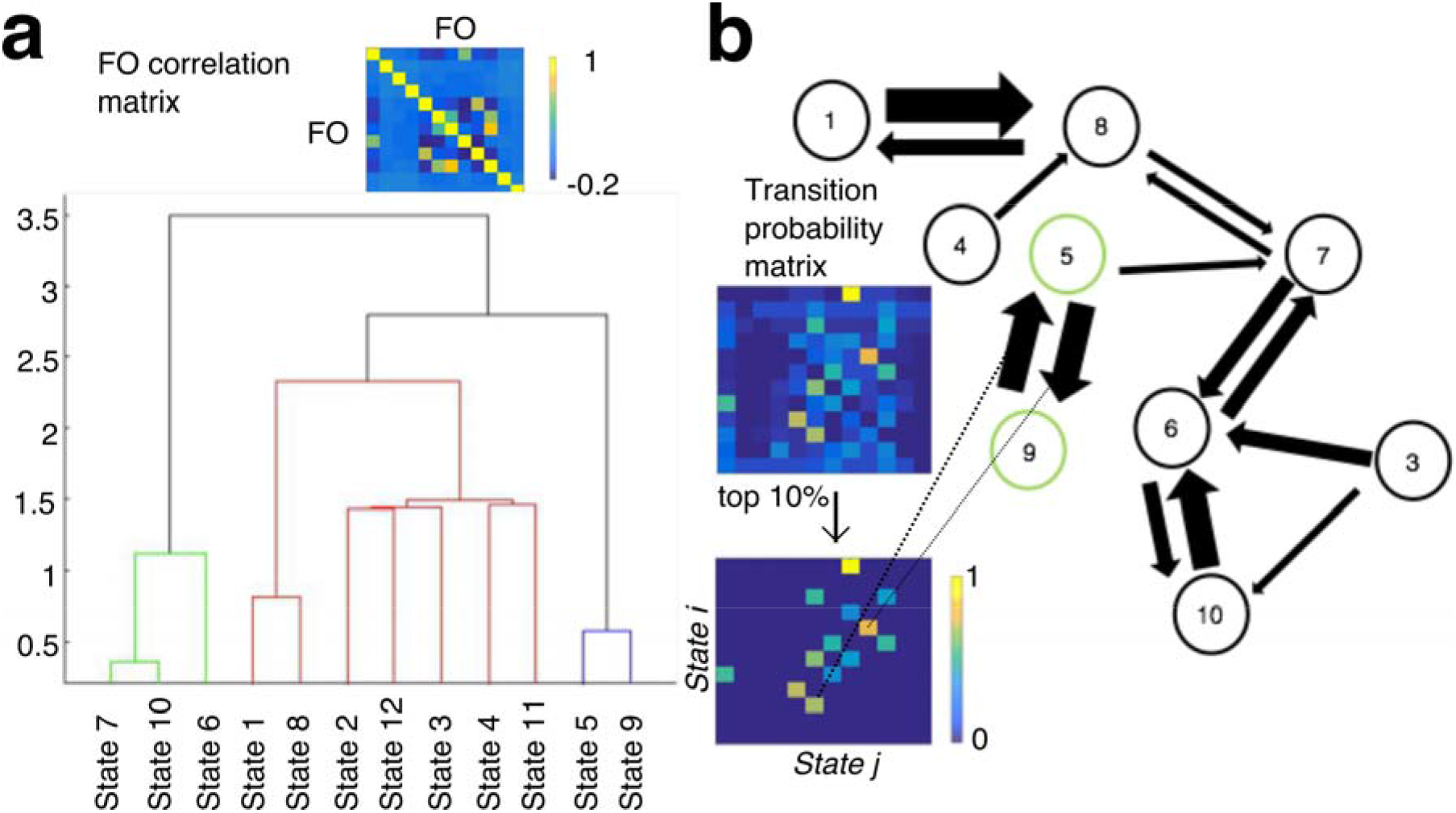
States 5 and 9 are temporally coherent. **a)** The fractional occupancy/FO correlation matrix shows how much each state is related to another in terms of their FOs across the patient cohort. Using a hierarchical clustering algorithm we identified that states 5 and 9 were clustered together at the lowest level of clustering. **b)** We also obtained the transition probability matrix for the patient cohort quantifying the probability of transitioning between different states. After obtaining the most significant transitions, we found that, when at states 5 and 9, the participants in the patient cohort would highly switch between these two rather than transitioning to another state.

## Discussion

TBI has been associated with a number of physical and cognitive sequelae; however, its association with depression has not yet been fully investigated with a siseable cohort. We used a novel method for quantifying brain dynamics in a patient cohort in order to investigate pending hypotheses regarding the constant recruitment of and switching between specific brain circuits. In summary we found a positive correlation between abnormal brain dynamics in patients and depression scores. Highly depressed patients showed abnormally high switching rate and FO in two states/networks spanning default mode, subcortical and cerebellar regions. These states were temporally coherent forming a metastate at which the depressive brain would visit.

The HMM was used to model brain activity as a sequence of distinct states/networks that the brain visits. This allowed us to uncover how much time each patient’s brain spent on each state (FO) as well as their switching rate. CCA revealed a positive relationship between brain dynamics and depression. Specifically, we identified a positive correlation between the FO and switching rate of specific states (states 5 and 9) and the depression score. In addition, comparison of the FO and switching rate between highly depressed and non-depressed patients revealed abnormally increased recruitment and switching in these states at high levels of depression. Spatial characterization of these two states resulted in identifying two distinct patterns. State 9 included parts of the DMN (mostly angular and posterior parts) whereas state 5 included subcortical, cerebellar and brainstem regions as well as anterior parts of the DMN. Regions of the DMN have been implicated in major depressive disorders (38). In an earlier (but smaller) study we reported altered functional connectivity in the subgenual ACC (part of the DMN) (10), which is linked to emotional processing as well as cognitive control via its reciprocal connections to DLFPC (12). Depression studies have also implicated subcortical regions and especially the thalamus. Specifically, they found reduced connectivity between the thalamus and DLPFC that might be associated with the decrease in the ability to execute attention-demanding tasks (10,39).

This result also aligns with previously proposed neuropsychological models of depression positing that negatively biased emotional perception is associated with increased recruitment of midline structures (for example the subgenual ACC). Thus it is possible that the high occupancy of these states reflects a ruminative, negatively oriented cognitive state in depressed individuals.

At the same time, the high switching rate of these two states supports the model proposing an inherent difficulty of these patients to disengage from increased rumination and negatively biased thinking (11). This inability could potentially find basis in dysfunctional bottom-up and top-down control between midline structures and the prefrontal cortex. Specifically, negatively biased emotional processing can lead to dysfunctional bottom up effects, for example from the subgenual ACC to DLPFC. In turn, DLPFC can exert dysfunctional top-down control via reciprocal connections to the ACC and the hippocampus (12). This dysfunctional top-down approach can be considered as “negative expectation” where depressed individuals act as if they expect negative information from their environment and are unable to exert cognitive control over it. Thus bottom up negative biases that are unsuccessfully kept under control by DLPFC can lead to an ongoing negative self-referential processing.

An additional point of discussion regarding the spatial characterization of our states is the inclusion of the brainstem. Although our spatial resolution precludes definite answers we believe that high occupancy and switching rate of these states in depressed patients could be connected to serotonergic deregulation. One possible hypothesis is that the previously proposed abnormal bottom-up, top-down loop is triggered by increased serotonin transmission via monoamine neurotransmitter projections from the brainstem to cortical and subcortical regions such as the ACC and the hippocampus. Furthermore, previous work has shown that serotonin reuptake inhibitors (served as anti-depressant drugs) have an effect on the stability of functional connectivity rendering it less volatile across time. Thus constant recruitment and high switching within the discovered states could potentially reflect a serotonin-specific effect on abnormal brain dynamics in depressed individuals (40). Further research, perhaps via pharmacological modulation, is required to verify this hypothesis.

We further discovered that these states related strongly to each other with similar temporal occupancies across participants and in terms of high inter-state transition probabilities. These findings suggest that these states behave as a high-level “metastate” with each participant constantly visiting and circulating within its sub-states. One interpretation of this result is that depression is associated with constant visiting of a self-referential “metastate” that reflects the tendency to dwell on past negative information (13).

The ultimate goal of TBI research is to manage and improve patients’ lives post TBI. While most rehabilitation techniques overlook changes in mood and neuropsychiatric symptoms, we believe it is important for modern methods to address these with the use of specific cognitive, emotional and motivational targets. In light of our findings and their relationship to the bottom-up, top-down model of depression, potential pharmacological therapies (selective serotonin reuptake inhibitors, SSRI’s) could target negative biases (bottom up) in the midline regions, whereas cognitive behavioural therapy (CBT) could target high level (top down) biases towards negative stimuli (for example TMS in the DLPFC) (11). Further imaging research in abnormal brain dynamics will provide more spatial specificity for these pharmacological and transcranial magnetic stimulation (TMS) targets. It is also worth noting a particular limitation of this study: despite our preliminary null finding on grey matter differences between groups, it is possible that the abnormal dynamics observed in these patients might be underpinned by other, non-observable with our current dataset changes in their underlying structural integrity (37). Further research is needed to investigate this relationship specifically by comparing our results to subjects with depression but no brain injury.

In conclusion, to date only a few studies exist that investigate depression post TBI and there is no clear consensus as to what are the neural correlates of depression in patients post TBI and what can be done to improve their daily lives. Refined approaches, such as the one presented here that quantify brain dynamics using resting-state fMRI, can contribute towards this direction by providing a marker of abnormal brain response and delineating the dysfunctional neural circuitry during depression. We argue that this is critical for understanding depression post TBI and deriving targeted therapies for improving the patients’ lives.

## Materials and Methods

### Patient cohort

A total of 81 subjects with TBI participated in this study. Patients with TBI were referred from the Addenbrooke’s Neurosciences Critical Care Unit Follow-Up Clinic and the Addenbrooke’s Traumatic Brain Injury Clinic.

Inclusion criteria included the presence of brain injury and age range between 16 and 60 years old. The exclusion criteria were (i) National Adult Reading Test < 70, (ii) Mini Mental State Exam < 23, (iii) left-handedness, (iv) contraindications for MRI scanning and pregnancy or nursing and (v) had a physical disability that could prevent them from completing the screening or scanning stages. All subjects gave written informed consent before participating in the study as approved by the Cambridgeshire Research Ethics Committee (Cambridge local Ethics committee reference number of approval LREC 97/290). TBI patients suffered from brain injury quantified by their Glasgow Outcome Scale (GOS) (20), Injury Severity Scale (ISS) (21), and Acute Physiology and Chronic Health Evaluation II (APACHE) (22) scores. Patients were at least 4 months post TBI (mean 15.20 ± 11.56 months) and were not receiving acute hospital interventions. Demographic and clinical characteristics of the patients are presented in Supplementary Table 1. Depressive symptomatology was evaluated with the Beck Depression Inventory (BDI) (23), a 21-item self-report scale measuring the emotional, cognitive, somatic, and motivational symptoms of depression. Each item is scored on a scale from zero to three, and total scores are calculated by summing the scores on all the items. Scores of ten or higher fall within the depressed range with scores from 10 to 16 indicating mild depression, scores from 17 to 29 moderate, and scores from 30 to 63 severe depression. Eventually patients were split in two groups in order to investigate differences in brain dynamics between patients with and without depression: the TBI no-depression group with BDI <10 and the TBI depression group with BDI >=10. The number of patients in each cohort was 52 and 29 respectively. There were no statistical differences in the IQ (two-tailed two-sample t-test, p=0.1761), sex (two-tailed two-sample t-test, p=0.2269) and age (two-tailed two-sample t-test, p=0.5061) between the two groups.

### MRI data acquisition

#### Acquisition parameters

Participants were scanned on a Siemens Trio 3-Tesla MR system (Siemens AG, Munich, Germany) at the Wolfson Brain Imaging Centre of Addenbrooke’s Hospital (Cambridge, UK). The imaging session started with a localizer followed by a high resolution T1 weighted, magnetization-prepared 180 degrees radio-frequency pulses and rapid gradient-echo (MPRAGE) structural scan (TR=2300ms, TE=2.98ms, TA=9.14min, flip angle=9°, field of view read=256mm, voxel size=1.0×1.0×1.0mm, slices=176). For the depressed cohort, fMRI assessment involved a resting state scan with a duration of 10 minutes (or 300 TRs out of which the first 5 were discarded due to signal stabilization issues) for which participants were instructed to not think of anything in particular and to keep their eyes closed. Image acquisition parameters for obtaining the functional data were as follows: EPI (echo-planar imaging) sequence with the following parameters: TR=2000ms, TE=30ms, flip angle=78°, field of view read=192 mm, voxel size=3.0×3.0×3.0 mm, slices per volume=32.

Preprocessing was done in SPM version 12 software (http://www.fil.ion.ucl.ac.uk/spm) and MATLAB Version 12a (Mathworks, Natick, MA) (http://www.mathworks.co.uk/products/matlab/) platforms. Functional images were slice time corrected, re-aligned, co-registered to the anatomical T1 weighted images and then normalised to MNI space using SPM’s unified segmentation method that segments brain tissue into gray matter, white matter and cerebrospinal fluid using tissue probability priors (24). Due to potential structural abnormalities caused by brain injury that can interfere with this process, we visually inspected the accuracy of each normalization. Normalised functional images were then smoothed with an 8 mm full width at the half maximum (FWHM) kernel.

We also obtained mean gray matter intensities in order to see if differences in brain dynamics between depressed and non-depressed TBI patients could be attributed to differences in structural abnormalities. For the gray matter analysis, the output tissue images were further refined using an iteration of a simple Markov Random Field that incorporates spatial prior information of the adjacent voxels into the segmentation estimation. The warped tissue types images were modulated using the Jacobian determinants obtained from the spatial normalization step. Finally the gray matter images were smoothed with a FWHM kernel of 8mm. Mean gray matter intensities used for comparison between the two groups were obtained as the mean of the gray matter smoothed images.

#### Whole-brain analysis

The smoothed normalised images were used to extract BOLD time courses from regions of interest spanning the entire brain. The regions of interest were defined using a combination of 91 cortical, 15 subcortical, and 26 cerebellar nodes coming from the Harvard-Oxford (25) and AAL parcellations (26). This atlas was provided with the CONN toolbox (27). In conjunction with this parcellation, we then used the CONN toolbox on the normalised smoothed images to extract time course from region of interest. An important preprocessing step when obtaining BOLD time courses is denoising as functional connectivity results have been shown to be sensitive to physiological noise (28). Nuisance regression was conducted using CompCor method (29). This method calculates the 5 first principal components of the white matter and cerebrospinal fluid signals and regresses them out from the gray matter signal. The regression also includes the motion parameters (as these come from the realignment process) and their first order derivatives. Despiking and linear detrending on the time courses was performed after regression. Finally, a high pass filtered [0.009, Inf] was applied to the denoised time courses.

Keeping in mind that the previous parcellation is defined mostly based on anatomical boundaries, analyses were also conducted using the Schaefer parcellation that parcellates the brain based on functional boundaries (30). This parcellation was derived based on clustering functional connectivity patterns thus associating brain areas with similar functional fingerprints. Thus to cover the entire brain, 400 cortical nodes were taken from the 400-node local-global parcellation 21 subcortical nodes were taken from the Harvard-Oxford parcellation and 22 cerebellar nodes were taken from the AAL atlas. Extraction of time courses was conducted using an identical process as the one with the previous parcellation.

### Relationship of brain dynamics to depression

#### Hidden Markov model

The HMM derives brain dynamics based on times series data obtained from the functional images as described in the previous section (**Fig. 1**). HMM assumes that the time series data are characterised by a number of states that the brain recurs at different times throughout the scanning period. At each time point t of brain activity, the observed time series data is modelled as a mixture of multivariate Gaussian distributions. Each one of these Gaussian distributions corresponds to a different state *k* and is described by first-order and second-order statistics (mean *μ_k_* and covariance *Σ_k_* respectively) that can be interpreted as the activity and functional connectivity of each state. Using notation, if *x_t_* describes the blood-oxygen-level-dependent (BOLD) data at each time point t, then we assume that the probability of being in state *k* is assumed to follow a multivariate Gaussian distribution

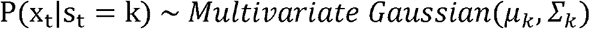

In turn, we wanted to model how transitions between states take place. The basic Markovian principle that describes the transition between states assumes that the probability of the data being in state *k* at time *t* relates only to the probability of being in state *l* at time *t* − 1. This can be described by the following equation

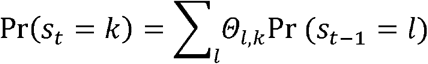

where *Θ_l,k_* is the transition probability from state *l* to state *k*. Taken together, HMM infers the Pr(*s_t_* = *k*) probabilities for each state *k* and time *t* (state time courses) as well as the transition probabilities *Θ_l,k_* and the statistics of each state (*μ_k_*, *Σ_k_*) that best describe the data. To make inference tractable a variational Bayes algorithm was used that works by minimizing the Kullback-Leibler divergence between the real and the modelled data.

The input time series data for HMM was the concatenated, denoised data as described in the whole-brain analysis section. We thus obtained a matrix of dimensions *number of images* × (*number of subjects* × *number of ROIs*) by concatenating the time series data from all subjects. Prior to concatenation, the participant-specific time series were standardised (centered and standard deviation = 1).

To reduce noise in the data matrix, a principal component analysis (PCA) dimensionality-reduction technique with #PCA components=25 was applied on the concatenated time courses (see comment on choosing PCA components in the *Supplementary Material*). The number of states was chosen as *k* = 12 (see comment on choosing the number of states in the Supplementary Material).

HMM results in state time courses representing the probability of each state being active in each time potion. Fractional occupancy (FO) of each state was defined as the proportion of time each patient spends on each state and quantified by its average activation probability over time. Switching rate was defined as the total (over time) amount of activation differences between consecutive time points for the states of interest. It was calculated by summing each state’s difference in activation probability between time t and time t+1 over time (and then summing over the states of interest). HMM also returns the mean activity *μ_k_* of each state. The HMMBOX MATLAB toolbox (www.robots.ox.ac.uk/~parg/software) was used to derive all the relevant HMM quantities used in this paper alongside custom-made scripts in MATLAB.

#### Correlation between dynamic functional connectivity and depression scores

After running the HMM we wanted to investigate whether the temporal characteristics (FO and switching rate) of the HMM states would relate to the participants’ depression scores. To do so we used a canonical correlation analysis (CCA). This analysis seeks maximal correlations (or modes) between two sets of variables, *X* and *Y*(31). CCA seeks linear combinations *U* and *V* of two sets of variables and (column vectors of dimension number of observations number of variables) such that they are maximally correlated. If a significant relationship exists between *U* and *V* then there exists a significant mode of co-variation between *X* and *Y* (CCA component). The variables *U* = *XA* and *V* = *YB* (more precisely in MATLAB these are defined as (*X* – *repmat*(*mean* (*X*), *N*, 1))* *A* and (*Y* – *repmat*(*mean*(*Y*)*, N*, 1)) * *B* where *N* =number of observations) are called canonical variables and correspond to linear combinations of variables of the sets *X* and *Y* respectively. In our case we wanted to see whether the combination of FO and switching rate of certain states would relate to the depression score. Thus we identified relevant *U* and *V* variables that corresponded to a significant CCA component between FO, switching rate and depression. *Post hoc* correlations of each column of *U* with each column of *X* (or *V* with each column of *Y*) quantified the contribution (and sign) of each of the variables to the CCA component. Statistical significance of each mode is obtained as described in the statistics method

#### Spatial and temporal characterization of states

After identifying relevant states using CCA, three separate analyses were conducted. First, we spatially characterised these states by mapping their mean activity *μ_k_* to the brain. Specifically, mean activity was projected to the original number of ROIs using the PCA loadings obtain from the PCA decomposition described previously. To help with interpretation of the results, the spatial maps were thresholded keeping the top 40% positively activated regions. Second, we assessed the uniqueness of their temporal characteristics by comparing the FO and switching rate in the TBI depression vs. TBI no-depression groups. Statistics were performed between these groups for the FOs of states 5 and 9 and their switching rates (see Statistics section). Third, we wanted to see if the temporal characteristics of these states were related i.e. if they were forming a metastate. To do so we used two methods. The first method considered the correlations between the FO of each state across all subjects. This 1 × *number of subjects* vector for state *i* was correlated with the corresponding vector for state *j*. Doing so for all pairs of states, we obtained a *number states* × *number of states* matrix where each (*i, j*) entry represented the extent to which the FOs for states (*i,j*) were correlated across subjects. In turn, we performed an agglomerative clustering analysis on the entries of this matrix. This analysis starts by classifying each data point as a separate cluster and progressively combines clusters of data at different hierarchical levels: similar data are clustered at a low level of hierarchy and less similar data are clustered at a higher level of hierarchy. We used the *linkage* function as implemented in MATLAB with default settings (method= ‘single’, distance= ‘euclidean’). The second method utilised the transition probabilities between each pair of states as obtained from the HMM. These can be represented as a matrix where each (*i, j*) entry stands for the probability of going from state *i* to state *j*. We used the top 10% entries in the probability transition matrix in order to identify the strongest transitions between states.

### Statistical analysis

No a priori power calculation took place although our sample size is larger than most MRI studies. Demographics between-group comparisons were performed using two sample t-tests. Before running any CCA analysis we wanted to select those states that had the highest FO. For getting confidence intervals on their fractional occupancy we calculated 10,000 permutations of the state time courses and we observed how many times their FO was less than the observed one. Confidence intervals were obtained using MATLAB’s binofit function (# FO < less than observed value, # permutation).

CCA modes were obtained from the MATLAB *canoncorr* function. Statistical significance was estimated as follows. We calculated 10,000 permutations of the rows of the *X* relative to *Y* and we re-calculated the CCA mode. Statistical significance was estimated using FWE correction across all modes computed (32). Additional validation included splitting the sample into train and test sets. CCA was calculated on the train set and applied to the test i.e. the matrices *A* and *B* were calculated in the train set and applied on the *X* and *Y* of the test set, We ran 1,000 iterations where we randomly chose 80% of the subjects as the train set and we obtained the correlation of the variables *U* and *V* in the respective test sets. We then reported the mean CCA mode across the 1,000 random splits. *Post hoc* correlation of each variable with the CCA mode provides a weight of how important each variable is to the CCA. Correlations were performed using the Spearman’s r and p value as obtained from the robust correlation toolbox provided by Pernet et al. (33) (http://sourceforge.net/projects/robustcorrtool/). Under the presence of not normally distributed data, correlation estimations using this toolbox provide more accurate false positive control without loss of power. All *post hoc* correlations were Bonferroni corrected for the number of correlations taking place. Finally, due to the fact that the HMM data (FO, switching rate) were not normally distributed, statistical comparisons between the depression and no depression groups were performed in non-parametric fashion using the two-tailed Wilcoxon rank sum test at alpha significance level 0.05 (34).

## Data Availability

All data and materials can be provided upon reasonable request to the authors

## Acknowledgments

We thank the clinicians at the Royal London Hospital Intensive Care Unit for their help with patient recruitment. We would also like to thank all the participants for their contribution to this study.

## Author contributions

IP, LML, BJS, DKM, and EAS conceived the study. IP, LML, BJS, DKM and EAS wrote the manuscript. IP, LML, and EAS carried out image preprocessing and statistical analyses. ELC, AM, JGO, JPC, VFN, DKM, and EAS assessed patients and were involved in patient recruitment and data acquisition.

## Data and materials availability

All data and materials can be provided upon reasonable request to the authors

## References

1. Maas AIR, Menon DK, Adelson PD, Andelic N, Bell MJ, Belli A, Bragge P, Brazinova A, Büki A, Chesnut RM, Citerio G, Coburn M, Cooper DJ, Crowder AT, Czeiter E, Czosnyka M, Diaz-Arrastia R, Dreier JP, Duhaime AC, Ercole A, van Essen TA, Feigin VL, Gao G, Giacino J, Gonzalez-Lara LE, Gruen RL, Gupta D, Hartings JA, Hill S, Jiang JY, Ketharanathan N, Kompanje EJO, Lanyon L, Laureys S, Lecky F, Levin H, Lingsma HF, Maegele M, Majdan M, Manley G, Marsteller J, Mascia L, McFadyen C, Mondello S, Newcombe V, Palotie A, Parizel PM, Peul W, Piercy J, Polinder S, Puybasset L, Rasmussen TE, Rossaint R, Smielewski P, Söderberg J, Stanworth SJ, Stein MB, von Steinbüchel N, Stewart W, Steyerberg EW, Stocchetti N, Synnot A, Te Ao B, Tenovuo O, Theadom A, Tibboel D, Videtta W, Wang KKW, Williams WH, Wilson L, Yaffe K, InTBIR Participants and Investigators. Traumatic brain injury: integrated approaches to improve prevention, clinical care, and research. Lancet Neurol 2017; 16:987–1048.

2. Whelan-Goodinson R, Ponsford J, Schönberger M. Validity of the hospital anxiety and depression scale to assess depression and anxiety following traumatic brain injury as compared with the structured clinical interview for DSM-IV. J Affect Disord 2009; 114:94–102

3. Levin HS, Brown SA, Song JX, McCauley SR, Boake C, Contant CF, Goodman H, Kotrla KJ. Depression and posttraumatic stress disorder at three months after mild to moderate traumatic brain injury. Journal of Clinical Experimental Neuropsychology 2017; 23:754–769.

4. Murphy FC, Michael A, Robbins TW, Sahakian BJ. Neuropsychological impairment in patients with major depressive disorder: the effects of feedback on task performance. Psychol Med 2003; 33:455–467.

5. Elliott R, Rubinsztein JS, Sahakian BJ, Dolan RJ. The neural basis of mood-congruent processing biases in depression. Archives of General Psychiatry 2002; 59:597–604.

6. Fedoroff JP, Starkstein SE, Forrester AW, Geisler FH, Jorge RE, Arndt SV, Robinson RG. Depression in patients with acute traumatic brain injury. Am J Psychiatry 1992; 149:918–23.

7. Dosenbach NU, Fair DA, Miezin FM, Cohen AL, Wenger KK, Dosenbach RA, Fox MD, Snyder AZ, Vincent JL, Raichle ME, Schlaggar BL, Petersen SE. Distinct brain networks for adaptive and stable task control in humans. Proc Natl Acad Sci USA 2007; 104:11073–11078.

8. Hamilton JP, Farmer M, Fogelman P, Gotlib IH (2015) Depressive rumination, the defaultmode network, and the dark matter of clinical neuroscience. Biol Psychiatry 78:224-230.

9. Andrews-Hanna JR, Smallwood J, Spreng RN. The default network and self-generated thought: component processes, dynamic control, and clinical relevance. Ann N Y Acad Sci 2014; 1316:29–52.

10. Moreno-López L, Sahakian BJ, Manktelow A, Menon DK, Stamatakis EA. Depression following traumatic brain injury: A functional connectivity perspective. Brain Inj 2016; 30:1319–1328.

11. Roiser JP, Elliott R, Sahakian BJ. Cognitive Mechanisms of Treatment in Depression. Neuropsychopharmacology. 2012; 37: 117-136.

12. Roiser JP, Sahakian BJ. Hot and cold cognition in depression. CNS Spectr 2013; 18:139–419.

13. Gotlib IH, Joormann J. Cognition and depression: current status and future directions. Annu Rev Clin Psychol 2010; 6:285–312.

14. Hutchison RM, Womelsdorf T, Allen EA, Bandettini PA, Calhoun VD, Corbetta M, Della Penna S, Duyn JH, Glover GH, Gonzalez-Castillo J, Handwerker DA, Keilholz S, Kiviniemi V, Leopold DA, de Pasquale F, Sporns O, Walter M, Chang C. Dynamic functional connectivity: promise, issues, and interpretations. Neuroimage 2013; 80:360–378.

15. Wise T, Marwood L, Perkins AM, Herane-Vives A, Joules R, Lythgoe DJ, Luh WM, Williams SCR, Young AH, Cleare AJ, Arnone D. Instability of default mode network connectivity in major depression: a two-sample confirmation study. Transl Psychiatry 2017; 7:e1105.

16. Kaiser RH, Whitfield-Gabrieli S, Dillon DG, Goer F, Beltzer M, Minkel J, Smoski M, Dichter G, Pizzagalli DA. Dynamic resting-state functional connectivity in major depression. Psychopharmacology 2016; 41:1822–1830.

17. Liégeois R, Laumann TO, Snyder AZ, Zhou J, Yeo BTT. Interpreting temporal fluctuations in resting-state functional connectivity MRI. Neuroimage 2017; 163:437–455.

18. Vidaurre D, Smith SM, Woolrich MW. Brain network dynamics are hierarchically organized in time. Proc Natl Acad Sci USA 2017; 114:12827–12832.

19. Vidaurre D, Abeysuriya R, Becker R, Quinn AJ, Alfaro-Almagro F, Smith SM, Woolrich MW. Discovering dynamic brain networks from big data in rest and task. Neuroimage 2017; S1053-8119(17)30548-7.

20. Teasdale G, Jennett B. Assessment of coma and impaired consciousness. A practical scale. Lancet 1974; 2:81–84.

21. Baker SP, O’Neill B, Haddon W, Long WB. The injury severity score: A method for describing patients with multiple injuries and evaluating emergency care. The Journal of Trauma 1974;14:187-196.

22. Knaus WA, Draper EA, Wagner DP, Zimmerman JE. APACHE II: A severity of disease classification system. Critical Care Medicine 1974;13:818-829.

23. Beck AT, Ward CH, Mendelson M, Mock J, Erbaugh J. An inventory for measuring depression. Archives of General Psychiatry 1961; 4:561–571.

24. Ashburner J, Friston KJ. Unified segmentation. Neuroimage 2005; 26:839–851.

25. Makris N, Schlerf JE, Hodge SM, Haselgrove C, Albaugh MD, Seidman LJ, Rauch SL, Harris G, Biederman J, Caviness VS Jr, Kennedy DN, Schmahmann JD. MRI-based surface-assisted parcellation of human cerebellar cortex: an anatomically specified method with estimate of reliability. Neuroimage 2005; 25:1146–1160.

26. Tzourio-Mazoyer N, Landeau B, Papathanassiou D, Crivello F, Etard O, Delcroix N, Mazoyer B, Joliot M. Automated anatomical labeling of activations in SPM using a macroscopic anatomical parcellation of the MNI MRI single-subject brain. Neuroimage 2002;15:273-289.

27. Whitfield-Gabrieli S, Nieto-Castanon A. Conn: a functional connectivity toolbox for correlated and anticorrelated brain networks. Brain Connect 2012; 2:125–141.

28. Duff EP, Makin T, Cottaar M, Smith SM, Woolrich MW. Disambiguating brain functional connectivity. Neuroimage 2018; 173:540–550.

29. Behzadi Y, Restom K, Liau J, Liu TT. A component based noise correction method (CompCor) for BOLD and perfusion based fMRI. Neuroimage 2007; 37:90–101.

30. Schaefer A, Kong R, Gordon EM, Laumann TO, Zuo XN, Holmes AJ, Eickhoff SB, Yeo BTT. Local-Global Parcellation of the Human Cerebral Cortex from Intrinsic Functional Connectivity MRI. Cereb Cortex 2018; 28:3095–3114

31. Hotelling H. Relations between two sets of variants. Biometrika 1936; 28:321–377.

32. Smith SM, Nichols TE, Vidaurre D, Winkler AM, Behrens TE, Glasser MF, Ugurbil K, Barch DM, Van Essen DC, Miller KL. A positive-negative mode of population covariation links brain connectivity, demographics and behavior. Nat Neurosci 2015; 18:1565–1567.

33. Pernet CR, Wilcox R, Rousselet GA. Robust correlation analyses: false positive and power validation using a new open source matlab toolbox. Front Psychol 2013; 3:606.

34. Hollander M, Wolfe DA. Nonparametric Statistical Methods. New York: Wiley-Interscience; 1999.

35. Greicius MD, Flores BH, Menon V, Glover GH, Solvason HB, Kenna H, Reiss AL, Schatzberg AF. Resting-state functional connectivity in major depression: abnormally increased contributions from subgenual cingulate cortex and thalamus. Biol Psychiatry 2007; 62:429–437.

36. Russo SJ, Nestler EJ. The brain reward circuitry in mood disorders. Nat Rev Neurosci 2013; 14:609–625.

37 Koolschijn PC, van Haren NE, Lensvelt-Mulders GJ, Hulshoff Pol HE, Kahn RS. Brain volume abnormalities in major depressive disorder: a meta-analysis of magnetic resonance imaging studies. Hum Brain Mapp 2009; 30:3719–3735.

38 Sheline YI, Price JL, Yan Z, Mintun MA. Resting-state functional MRI in depression unmasks increased connectivity between networks via the dorsal nexus. Proc Natl Acad Sci USA 2010; 107:11020–11025.

39. Fales CL, Barch DM, Rundle MM, Mintun MA, Snyder AZ, Cohen JD, Mathews J, Sheline YI. Altered emotional interference processing in affective and cognitive-control brain circuitry in major depression. Biol Psychiatry 2008; 63:377–384.

40. Nemeroff CB, Owens MJ. The role of serotonin in the pathophysiology of depression: as important as ever. Clin Chem 2008; 55:1578–1579.

